# Near-Fibre Electromyography

**DOI:** 10.1101/2020.10.07.20208348

**Authors:** Mathew Piasecki, Oscar Garnés C. Estruch, Daniel W Stashuk

**Affiliations:** Clinical, Metabolic and Molecular Physiology, MRC-Versus Arthritis Centre for Musculoskeletal Ageing Research, National Institute for Health Research (NIHR) Nottingham Biomedical Research Centre, University of Nottingham, Nottingham, United Kingdom; Department of Physical Medicine and Rehabilitation - Clinical Neurophysiology, Jiménez Díaz Foundation University Hospital, Madrid, Spain; Department of Systems Design Engineering, University of Waterloo, Ontario, Canada

**Keywords:** near fibre electromyography, near fibre motor unit potential, quantitative electromyography

## Abstract

Near fibre electromyography (NFEMG) is the use of specifically high-pass filtered motor unit potential (MUPs) (i.e. near fibre MUPs (NFMs)) extracted from needle-detected EMG signals for the examination of changes in motor unit (MU) morphology and electrophysiology caused by neuromuscular disorders or ageing. The concepts of NFEMG, the parameters used, including NFM duration and dispersion, which relates to fibre diameter variability and/or endplate scatter, and a new measure of neuromuscular junction transmission (NMJ) instability, NFM segment jitter, and the methods for obtaining their values are explained. Evaluations using simulated needle-detected EMG data and exemplary human data are presented, described and discussed. The data presented demonstrate the ability of using NFEMG parameters to detect changes in MU fibre diameter variability, end plate scatter, and neuromuscular transmission time variability. These changes can be detected prior to alterations of MU size, numbers or muscle recruitment patterns.

## 1.0 Introduction

A motor unit (MU) is comprised of a somatic motor neuron (MN), its axon and distal axonal branches, neuromuscular junctions (NMJs) and associated skeletal muscle fibres. Neuromuscular disorders can affect somatic MNs, their axons, axonal branches, NMJs and/or associated muscle fibres. Anatomically neuromuscular disorders can cause loss of somatic MNs, axonal demyelination, and MU remodelling; fibre denervation and the sprouting of new axonal branches and the formation of new NMJs, acting to minimise muscle fibre loss; and fibre atrophy and/or hypertrophy. Electrophysiologically, neuromuscular disorders can cause NMJ transmission delay, increased NMJ transmission time variability and/or NMJ transmission failure (Daube 2000). An effective way of detecting the anatomical and electrophysiological effects of a neuromuscular disorder, and therefore its presence and extent, is to examine electrical signals generated by nerves and muscles. With respect to an affected muscle, these anatomical and electrophysiological alterations can be inferred by characterizing its electrophysiology via the detection and analysis of electromyographic signals.

The discharge/activation of a MN creates a trans-axolemmal or axonal action potential (AAP), which propagates along its axon and each axonal branch to initiate neuromuscular transmission to its associated muscle fibre. Successful neuromuscular transmission results in the generation of propagating trans-sarcolemmal or muscle fibre action potentials (MFAPs) and coordinated contraction of the sarcomeres of the associated muscle fibre. As such, the MU is the fundamental element of muscle force generation (Heckman and Enoka 2012).

Extracellular electrodes placed in or on a skeletal muscle can detect the time changing electric potentials created by the propagating MFAPs of a MU. The time changing electric potential created by the propagating MFAPs of a single muscle fibre can be considered a muscle fibre potential (MFP) while the summation of the MFPs created by the fibres of a MU is called a motor unit potential (MUP). For a healthy MU, each discharge of a MN will result in the generation of a MFAP in each fibre of the MU. As such, MUPs represent the summation of the MFPs of all of the MU fibres.

During the activation of a healthy muscle, populations of MUs are recruited and independently discharge at pseudo-regular firing times to generate a desired level of muscle force. With simple level or window triggering or more sophisticated pattern recognition methods, MUP trains (MUPTs) (i.e. sets of consecutively detected MUPs generated by the same MU) can be extracted from a recorded EMG signal. The number of trains extracted can be indicative of the level of MU recruitment. The occurrence times of the MUPs within MUPTs can be indicative of MU discharge patterns. Both of these are related to a combination of muscle anatomy and electrophysiology. For known levels of activation, the number of MUs recruited, their discharge patterns and anatomical characteristics can be used to infer the presence and extent of a neuromuscular disorder.

MU recruitment and firing patterns can be used to infer the relative numbers and sizes of MUs in a muscle (Sonoo 2002). A typical MUP within a train or a calculated MUP template can be used to infer MU anatomical features, such as size and the extent of terminal axonal branching. MUP shape instability within a train can be used to infer electrophysiological features, such as terminal AAP and/or MFAP conduction velocity variability in combination with NMJ transmission time variability. NMJ transmission time variability is often much greater than AAP and/or MFAP conduction velocity variability and therefore greater MUP instability largely reflects NMJ transmission time variability.

More specifically, standard clinical EMG examinations are now, almost exclusively, based on signals detected using concentric needle electrodes. During low-level muscle activation, MUPTs are extracted using triggering or pattern recognition techniques. MU recruitment is qualitatively –i.e. subjectively-assessed at increasing levels of force, whereas discharge patterns are usually not considered. A representative or template MUP is used to infer MU size based on MUP duration, amplitude and/or area. Increased MUP size is expected to be related to axonal sprouting which is in turn related to re-innervation of denervated muscle fibres and indicative of MN loss. Reduced MUP size is expected to be related to fibre loss, atrophy or NMJ blocking. Axonal sprouting and fibre atrophy or hypertrophy are indirectly assessed using the number of phases and/or the number of turns in the representative or template MUP. The formation of new NMJs and NMJ transmission delay, NMJ transmission time variability and/or NMJ transmission failure are assessed using either separate SFEMG examination techniques (which are more accurate though time consuming) or using a triggering technique and high-pass filtered EMG signals at low levels of contraction to evaluate MUP complexity and morphological instability (i.e. jiggle).

Specific aspects of neuromuscular disease could be better detected if signals better reflecting the anatomical and electrophysiological effects of disease on individual muscle fibres could be assessed. In general, it is not possible to detect only the MFP created by the MFAP of a single muscle fibre. However, as the characteristics of a MFP generated by a muscle fibre are dependent on fibre diameter and its radial distance to the electrode detection surface, as radial distance increases, the amplitude and high frequency content of a detected MFP decreases. This means that MFP contributions to a MUP from distant fibres will be of lower amplitude and have a greater proportion of lower frequency energy. Conversely, MFP contributions from near fibres (NFs) will be of higher amplitude and have a greater proportion of higher frequency energy. Electrodes with smaller detection surface area increase the disparity of the radial distances across the fibres of MU and, for a given electrode position relative to the fibres of a MU, allow some fibres to be effectively near the electrode and some to be distant. For a suitably positioned electrode with small detection surface area, the number of muscle fibres with small radial distance (i.e. the number of NFs) can be small and the detected MUP will be primarily composed of NF MFPs and significant NF MFP contributions may be detected. As suggested by Payan (1978) the use of high-pass filtering enhances the effects of radial distance, and in effect, for the same detection surface area, reduces the number of NFs and increases the chance of detecting significant NF MFP contributions. This is essentially the basis of single-fibre electromyography (SFEMG) whether using a traditional single-fibre or concentric needle electrode (Sanders & Stålberg, 1996; Stålberg, 2012). Because of the enhancement of NF contributions relative to distant fibre contributions, a suitably high-pass filtered concentric-needle detected MUP is called a NF MUP (NFM).

Near fibre electromyography (NFEMG) is the study of NFMs for the assessment of the sprouting of new unmyelinated axonal branches and the formation of new NMJs, NMJ transmission delay, NMJ transmission time variability or NMJ transmission failure and muscle fibre loss, atrophy or hypertrophy. Several previous studies (Allen et al. 2015; Hourigan et al. 2015; Piasecki et al. 2016b, 2016a, 2020; Power et al. 2016; Gilmore et al. 2017; Estruch and Stashuk 2019; Estruch et al. 2019) have incorporated NFEMG parameters to demonstrate the effects of specific diseases or ageing on individual MUs. The focus of this paper is to utilise MUPs and NFMs extracted from simulated EMG signals and those obtained from human muscles to explicitly describe and evaluate a range of NFEMG methods and parameters used to investigate neuromuscular alterations, including a new measure of NMJ instability, NFM segment jitter. Methods for MUPT extraction, high-pass filtering, MUP and NFM contamination reduction and parameter calculation are outlined.

## 2.0 Methods

### 2.1 Generation of Simulated Motor Unit Data

A commercially available application for creating simulated needle-detected EMG signals was used to create sets of signals detected using an electrode of known geometry and generated by MUs for which the numbers of fibres, their sizes, and their radial positions and NMJ axial positions, relative to the detection surface of the electrode, are known (Stålberg & Karlsson, 2001). The simulated EMG signals used are simple compared to real EMG data, lacking external 50/60Hz noise and movement artifacts, however they provide a basis from which useful evaluations of MUP/NFM parameters can be evaluated.

The simulation of an EMG signal starts by modelling MUs which comprise a muscle in which the signals are detected. In this work, a muscle comprised of four MUs was modelled. For each MU, the number of fibres, their fibre diameter and NMJ axial location distribution and their NMJ transmission time variability (i.e. jitter), as well as the expected diameter of the MU territory was selected. Firing pattern characteristics of each MU were also selected. To evaluate the separate effects of fibre diameter variability, end plate scatter, MU remodelling and jitter on MUP and NFM template and instability parameter values, specific muscle/EMG detection scenarios were modelled. Except for specific changes related to the scenarios described below, all muscles were simulated with default properties and signals were simulated as being recorded using the enlarged uptake area of a CNEMG electrode positioned axially 20 mm from the center of the endplate region, and randomly positioned radially within the territories of the simulated MUs. For each scenario, ten EMG signals each containing 4 MUPTs were simulated for 10 unique radial needle positions. Forty MUPTs per scenario were then extracted from the EMG signals using DQEMG algorithms.

### 2.2 Exemplary human data

Exemplary data is presented through three examples (Figures 5 to 7), corresponding to a control, neurogenic and myopathic subject. Muscles sampled were the gastrocnemius, gastrocnemius, extensor digitorum communis and vastus lateralis, and deltoid respectively, as detailed in section 3.5.

Data for all three cases were collected through retrospective review and were analysed using DQEMG® with the approval of the local ethics committee (Biomedical Research Institute, Jiménez Díaz Foundation University Hospital; code EO181-20_FJD). Diagnoses were those made by the examining neurophysiologist (OGE) at the time of the study, based on the results of the clinical and neurophysiological examination. EMG signals were acquired during a sustained mild effort protocol of 10 sec duration as part of the routine neurophysiological examination. Intramuscular signals were detected with a standard concentric needle electrode (38 x 0.45 mm (1.5” x 26G) Neuroline Concentric; Ambu®, Denmark). Raw EMG signals were bandpass filtered (20 Hz - 10 kHz) and stored using a KeyPoint.Net 3.22® device (Alpine Biomed, USA).

### 2.3 MUPT Extraction

To obtain representative as well as shape instability MUP/NFM information a MUPT must be extracted from a recorded EMG signal from which a representative/template MUP/NFM can be selected or estimated and across which MUP/NFM shape instability can be measured. MUPTs can be extracted using level or window trigger or pattern recognition algorithms. In this work, MUPTs were extracted from simulated or real concentric needle-detected EMG signals using the algorithms contained in DQEMG (Stashuk 1999a). These algorithms execute the following steps for MUPT extraction and analysis: 1) MUP detection, 2) initial MUP clustering, 3) supervised MUP classification, 4) MUPT splitting and merging, 5) extracted MUPT characterization. During MUPT extraction, the current DQEMG algorithms specifically account for MUP instability within a MUPT.

### 2.4 Choice of High-Pass Filter

To increase the effects of the radial distances to muscle fibres on detected MUPs and thus enhance contributions of near fibres to detected MUPs, the detected EMG signals are high-pass filtered. For conventional SFEMG analysis signals are usually filtered using a Butterworth high-pass filter with a 500 or 1000 Hz cut-off frequency. Earlier work (Stashuk 1999b), has demonstrated that the use of low-pass-double-differentiation (LPDD) filters (Usui and Amidror 1982; McGill et al. 1985) have similar amplitude responses as traditionally-used Butterworth filters, but because of different phase responses create MUPs in which it is easier to detect the contribution of single muscle fibres (i.e. MFPs). Therefore, in this work, the NFMs analyzed have been created by LPDD filtering concentric-needle-detected MUPs.

### 2.5 Definition of representative/template MUP and NFM parameters

MUP Area - integral of the absolute value of the MUP values between the onset and end markers, times the sampling time interval, in μVms.

MUP Duration - duration between the onset and end markers, in ms.

Turns – number of significant slope reversals within the MUP duration (height > 20 μv)

NFM Area - integral of the absolute value of the NFM values between the onset and end markers, times the sampling time interval, in kV/s^2^ms.

NFM Duration – duration between the onset and end markers of the NF MUP, in ms.

NF Count – number of fibre contributions detected within the NFM duration (i.e. significant slope reversals (height > 5*NF baseline RMS) with similar rising and falling slopes)

NF Dispersion – time between the first and last detected fibre contribution, in ms

### 2.6 Definition of MUPT parameters

One objective of analyzing the MUPs comprising a MUPT is to characterize the discharge pattern of the associated MN using the occurrence or detection times of the composite MUPs. An additional objective is to assess NMJ transmission time variability and/or AAP and MFAP conduction time variability. The former of these is the focus of SFEMG, in which a fibre pair (FP) jitter measurement is obtained when a pair of fibre contributions are repeatedly detected in successive MUPs across a MUPT, and the variance of their inter potential intervals (IPIs) is used to characterize the NMJ transmission time variability of the fibre pair. To obtain a measure related to the NMJ transmission time variability of a larger number of NMJs of a MU, statistics related to the instability of the shapes of the MUPs of a MUPT, such a jiggle, have been developed (Stålberg & Sonoo, 1994). In this work, MUP jiggle has been measured using the MUPs of MUPTs as well as NFM jiggle using LPDD filtered MUPs of MUPTs.

Jiggle values, and those of other similar global-MUP-shape based statistics (Stålberg & Sonoo, 1994), represent a larger number of NMJs of a MU than do fibre pair jitter measurements, are dependent on shape changes and therefore the relative sizes of MUP segments and not purely their temporal variability, and have normalized units which indirectly represent NMJ transmission time variability. A new statistic called NFM segment jitter (NFM SJ) is introduced in this work, which represents a larger number of NMJs of a MU than do fibre pair jitter measurements, is independent of the relative sizes of MUP segments and more dependent on their temporal variability and is measured in μs, to more directly represent NMJ transmission time variability.

To accurately assess NMJ transmission and/or AP conduction velocity variability using extracted MUPTs, the MUPs of each MUPT must first be aligned to each other. In this work, MUPs within a train are aligned during the process of MUPT extraction using methods similar to the correlation maximization method of Campos et al (2000).

#### 2.6.1 Calculation of NFM Segment Jitter (NFM_SJ)

Given a set of N, NFMs comprising a MUPT, let NFM_i_ represent the i^th^ NFM of the MUPT. Each NFM_i_ is a set of consecutive sample values.

Each NFM_i_ of the MUPT is divided into S_i_ segments.

A NFM segment is an interval of a NFM over which the absolute change in the amplitude of the NFM amplitude is equal to a threshold amount, segmentHeight.

NFM_ij_ is the j^th^ segment of NFM_i_.

NFM_ij_ is a set of consecutive samples from NFM_i_ such that NFM_ij1_ is the starting index in NFM_i_ of the j^th^ segment and NFM_ij2_ is the ending index in NFM_i_ of the j^th^ segment.

Therefore, |NFM_ij1_ – NFM_ij2_| = segmentHeight

The segments of a NFM are contiguous.

Therefore, NFM_i(j+1)1_ = NFM_ij2_ + 1

WeightedShiftSum = 0

SumOfWeights = 0

~~~
For i = 1 to i = N
 For j = 1 to j = S_i_
  If (i < N)
   Calculate the optimal shift s_ij_ between segment NFM_ij_ and NFM_i+1_
   by successively shifting NFM_ij_ relative to NFM_i+1_ to maximize their shape similarity w_ij_
   WeightedShiftSum = WeightedShiftSum + s_ij_*w_ij_
   SumOfWeights = SumOfWeights + w_ij_
  If (i > 1)
   Calculate the optimal shift s_ij_ between segment NFM_ij_ and NFM_i-1_
   by successively shifting NFM_ij_ relative to NFM_i-1_ to maximize their shape similarity w_ij_
   WeightedShiftSum = WeightedShiftSum + s_ij_*w_ij_
   SumOfWeights = SumOfWeights + w_ij_
NFM Segment jitter = WeightedShiftSum / SumOfWeights
~~~

In this work, the value of segmentHeight was the greater of 0.10% of the NFM template peak to peak voltage and 5 times the baseline RMS.

The value of s_ij_ was constrained to be in the range between −200 μs to +200 μs.

### 2.7 Definition and Selection of Isolated MUPs/NFMs

To accurately assess NMJ transmission and/or AP conduction velocity variability using detected MUPs/NFMs it is also essential that the MUPs/NFMs used represent the activation of a single MU and not be contaminated by the activity of other MUs. Such MUPs/NFMs, can be characterized as isolated MUPs/NFMs. To select isolated MUPs/NFMs within a MUPT, NFMs contaminated with contributions of other MUs need to be excluded. MUP/NFM contamination will cause a segment of a MUP/NFM to have significantly different values than corresponding segments of previous and subsequent MUPs/NFMs. Detecting these significant differences must take into account the inherent instability of the shapes of the NFMs within the train as well as any trend in NFM shape across the train due to needle movement. MUP/NFM instability measures are calculated over the NFM duration of the representative or template NFM. Therefore, an isolated NFM only needs to be contamination free across the NFM duration interval.

#### 2.7.1 Selecting Isolated NFMs

An isolated NFM is not significantly contaminated by the activity of other MUs.

A set of isolated NFMs allow the estimation of the true electrophysiological instability of the NFMs generated by a MU.

Given a set of N, NFMs comprising a MUPT, let NFM_i_ represent the i^th^ NFM of the MUPT.

Each NFM_i_ is a set of consecutive sample values.

Let NFM_t be the NF MUP template used to represent the N, NFMs of the MUPT.

The NFM_t of the MUPT is divided into S_t_ segments.

A NFM_t segment is an interval of the NFM_t over which the absolute change in the amplitude of the NFM_t amplitude is equal to a threshold amount, segmentHeight.

NFM_t_j_ is the j^th^ segment of the NFM_t.

NFM_t_j_ is a set of consecutive samples from the NFM_t such that NFM_t_j1_ is the starting index in the NFM_t of the j^th^ segment and NFM_t_j2_ is the ending index in the NFM_t of the j^th^ segment.

Therefore, |NFM_t_j1_ – NFM_t_j2_| = segmentHeight

The segments of the NFM_t are contiguous.

Therefore, NFM_t_(j+1)1_ = NFM_t_j2_ + 1

Each of the N NFMs in the MUPT are positioned as rows in a two dimensional array with N rows and k columns, where k is the number of samples in NFM_t.

Each of the N NFMs in the MUPT are segmentally aligned with the respective S_t_ segments of NFM_t to produce k columns of aligned NFM values.

Across each of the k aligned NFM value columns an average mean absolute consecutive difference (MACD) is calculated, with MACD_j_ being the value for the j^th^ column.

A NFM is considered isolated if for each of its k samples the average absolute deviation of m consecutive samples, centered at k, from m corresponding samples of the previous isolated NFM and from m corresponding samples of the NFM template are less than 10*MACD_k_, where m corresponds to the number of samples required to span 100 μs.

Having a vector of MACD values allows the level of expected variation (i.e. the amount of acceptable contamination) to be different for each time point across the set of NFMs with in the MUPT. Therefore, the threshold values used as criteria for significant contamination in the baseline segments of the MUPT will be more restrictive (have smaller values) that the threshold values used during the higher energy segments. This in turn, allows the shape variability across the NFMs of the MUPT to be accurately measured. The alignment of portions of the NFMs with the NFM template allow for more accurate estimation of the noise present.

### 2.8 Statistical Analysis

All statistical analysis was completed using STATA (v.15). All parameters within each scenario were assessed across levels of severity (i.e. increasing fibre diameter variability) using multi-level mixed effects linear regression models, with each needle position a fixed factor. Beta (β) coefficients and 95% confidence intervals are reported. Significance was accepted at p<0.05.

## 3.0 Results

### 3.1 Increasing fibre diameter variability

Four sets of 40 MUPTs were extracted from EMG signals recorded from muscles with MUs having default mean fibre diameters (50 μm) with diameter standard deviations of 1, 5, 10 and 15 μm. Figure 1 panels A to C show MUP and NFM size parameters across increases in fibre diameter variability. Increased MFP temporal dispersion associated with increased fibre diameter variability decreases MUP area (Figure 1A) and increases MUP duration (Figure 1C), but does not affect NFM area (Figure 1B).

**Figure 1.**
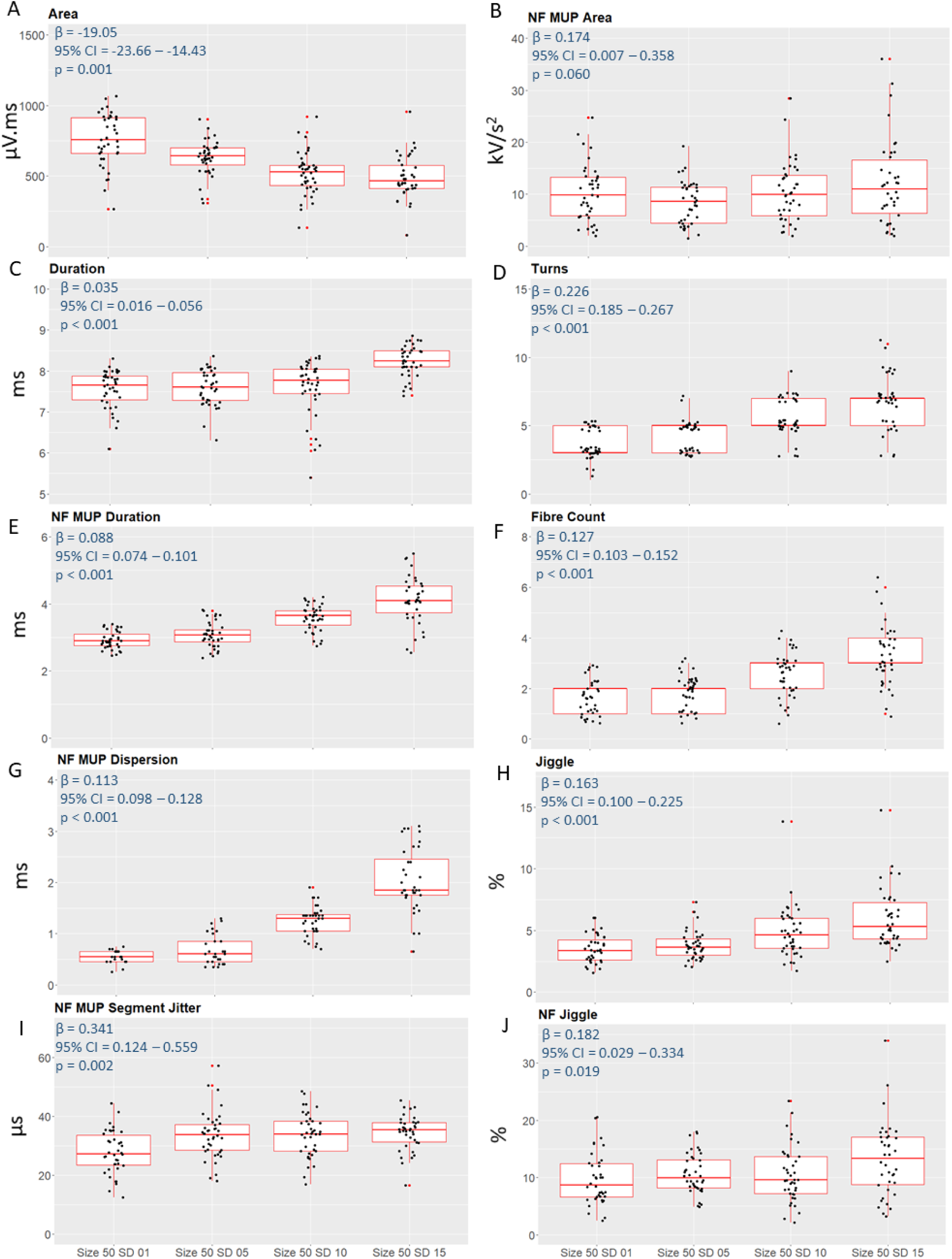
Motor unit potential and near fibre motor unit potential parameters with increases in fibre diameter variability. Beta coefficients, 95% confidence intervals and p values from multi level mixed effects linear regression shown inset for each parameter.

Figure 1 panels D and F show number of MUP turns and NF count values across increases in fibre diameter variability, respectively. All of these complexity related measures increase with the increased MFP temporal dispersion associated with increased fibre diameter variability. With increased MFP dispersion, increased numbers of underlying MFP contributions become evident/detectable. Figure 1 panels E and G show NFM duration and NF dispersion across increases in fibre diameter variability, respectively. Increases in MFP temporal dispersion are clearly and consistently reflected in all of these measures. Figure 1 panels H and J show MUP and NFM jiggle values, while panel I shows NFM segment jitter values across increases in fibre diameter variability. Even though the modelled NMJ jitter was constant, across the increasing levels of modelled fibre diameter variability, increased average MFP dispersion caused increased shape instability. This caused more consistent increases in MUP versus NFM jiggle, evidenced by the larger relative β coefficient (β/CI) and for MUP jiggle. The temporally-based NFM segment jitter values show moderate increases as indicated by the small relative β value.

### 3.2 Increasing NMJ axial location variability

Five sets of 40 MUPTs were extracted from EMG signals recorded from muscles with MUs having constant fibre size, and mean axial endplate locations of 0 mm with endplate location standard deviations of 1, 2, 5, 7, and 10 mm. Figure 2 panels A to C show MUP and NFM size parameters across increases in end plate scatter. As with increasing fibre diameter variability, increased MFP temporal dispersion associated with increased end plate scatter clearly decreases MUP area (Figure 2A) and increases MUP duration (Figure 2C), but only moderately increases NFM area (Figure 2B).

**Figure 2.**
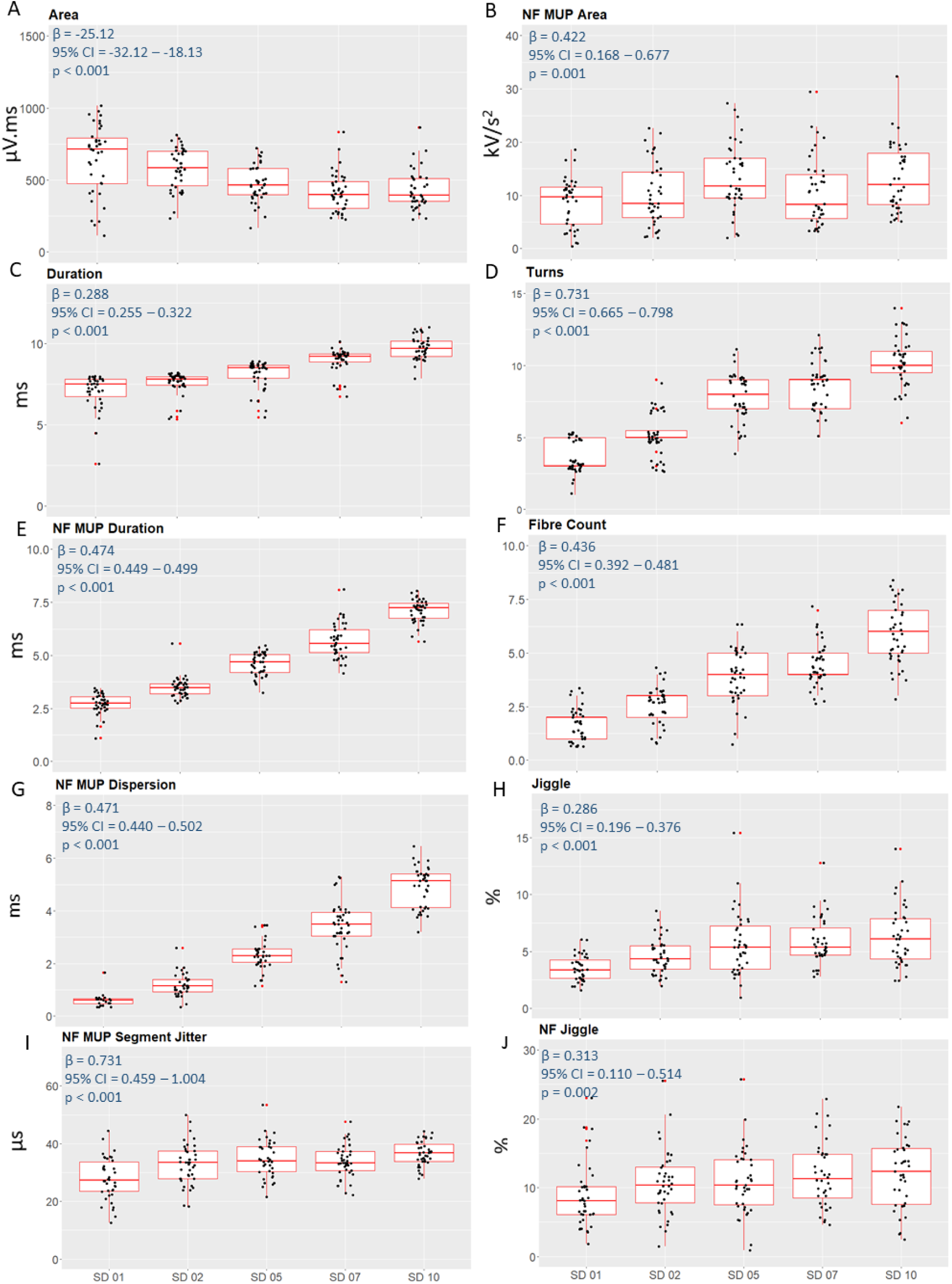
Motor unit potential and near fibre motor unit potential parameters with increases in end plate scatter. Beta coefficients, 95% confidence intervals and p values from multi level mixed effects linear regression shown inset for each parameter.

Figure 2 panels D and F show number of MUP turns and NF count values across increases in end plate scatter, respectively. All of these complexity related measures increased with the increased MFP temporal dispersion associated with increased end plate scatter. The increases in numbers of turns and NF count were greater than for the study of increased fibre diameter variability (range of mean turns values 4-10 versus 4-8; range of mean NF count values 2-6 versus 2-3). Figure 2 panels E and G show NFM duration and NF dispersion values across increases in end plate scatter, respectively. Increases in MFP temporal dispersion are clearly and consistently reflected in all of these measures. As with the complexity measures, the NFM duration, and NF dispersion values were about greater than for the study of increased fibre diameter variability (range of mean NFM duration values 2.5-7.5 versus 2.5-4; range of mean NF dispersion values 0.5-4.5 versus 0.5-2). Figure 2 panels H and J show MUP and NFM jiggle values, while panel I shows NFM segment jitter values across increases in end plate scatter. Even though the modelled NMJ jitter was constant, across the increasing levels of modelled end plate scatter, increased average MFP dispersion lead to increased shape instability. This caused more consistent increases in MUP versus NFM jiggle, evidenced by the larger relative β coefficient (β/CI) for MUP jiggle. The temporally-based NFM segment jitter values show clear increases as indicated by the large relative β value.

### 3.3 Simulated MU diminution and expansion

To consider a simple model of partial/distal denervation and reinnervation with fibre atrophy and hypertrophy, four sets of 40 MUPTs related to 4 degrees of involvement were extracted. The MUs were simulated to have characteristic as outlined in Table 1 below. Of the 4 simulated MUs for each muscle, 2 have ‘denervated’ and 2 have ‘compensated/reinnervated’ characteristics. Where fibre number is increased, so is fibre diameter variability and endplate scatter.

**Table 1.**
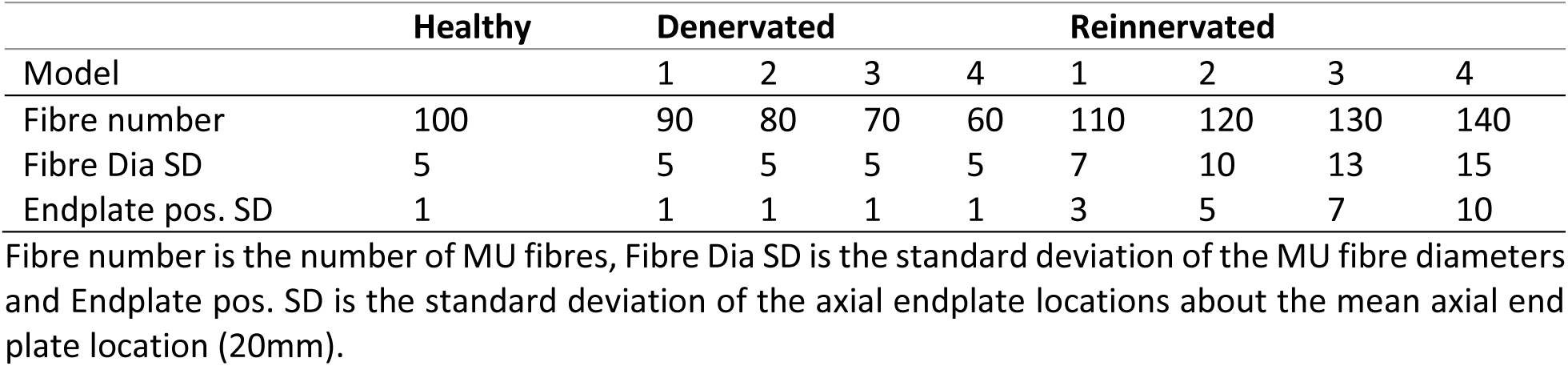
Features of remodelled motor units

|  | Healthy | Denervated |  |  |  | Reinnervated |  |  |  |
| --- | --- | --- | --- | --- | --- | --- | --- | --- | --- |
| Model |  | 1 | 2 | 3 | 4 | 1 | 2 | 3 | 4 |
| Fibre number | 100 | 90 | 80 | 70 | 60 | 110 | 120 | 130 | 140 |
| Fibre Dia SD | 5 | 5 | 5 | 5 | 5 | 7 | 10 | 13 | 15 |
| Endplate pos. SD | 1 | 1 | 1 | 1 | 1 | 3 | 5 | 7 | 10 |

Fibre number is the number of MU fibres, Fibre Dia SD is the standard deviation of the MU fibre diameters and Endplate pos. SD is the standard deviation of the axial endplate locations about the mean axial endplate location (20mm).

Figure 3 panel A shows MUP area decreasing with denervation and remaining unchanged with reinnervation relative to the default model. Only at the highest level of involvement simulated do reinnervated MUs have increased MUP area relative to denervated MUs. For denervated MUs, the decreases in MUP area are not due to increased temporal dispersion of the MFPs comprising the simulated MUPs but only the reduced numbers of MU fibres. For the reinnervated MUs, the increased temporal MFP dispersion, caused by increased fibre diameter variability and increased end plate scatter, combined with increased numbers of MU fibres result in no clear trend in MUP area. MUP duration however, shown in Figure 3 panel C, clearly decreases and increases for denervated and reinnervated MUs, respectively, as expected. NFM area, shown in Figure 3 panel B, did not change with denervation. With reinnervation, NFM area was increased with respect to the default and denervated models, but with increased variability no trend with reinnervation was evident. Figure 3 panels D and F do not show any trends in the numbers of turns and NF count values with increased denervation. The constant fibre diameter variability and end plate scatter modelled across the levels of increased denervation modelled resulted in no changes to these complexity measures despite the reduced numbers of MU fibres. These panels do show increasing trends in the numbers of turns and NF count values with increased reinnervation. The increased fibre diameter variability and end plate scatter modelled across the levels of increased reinnervation modelled resulted in clear changes to these complexity measures despite the increased numbers of MU fibres.

**Figure 3.**
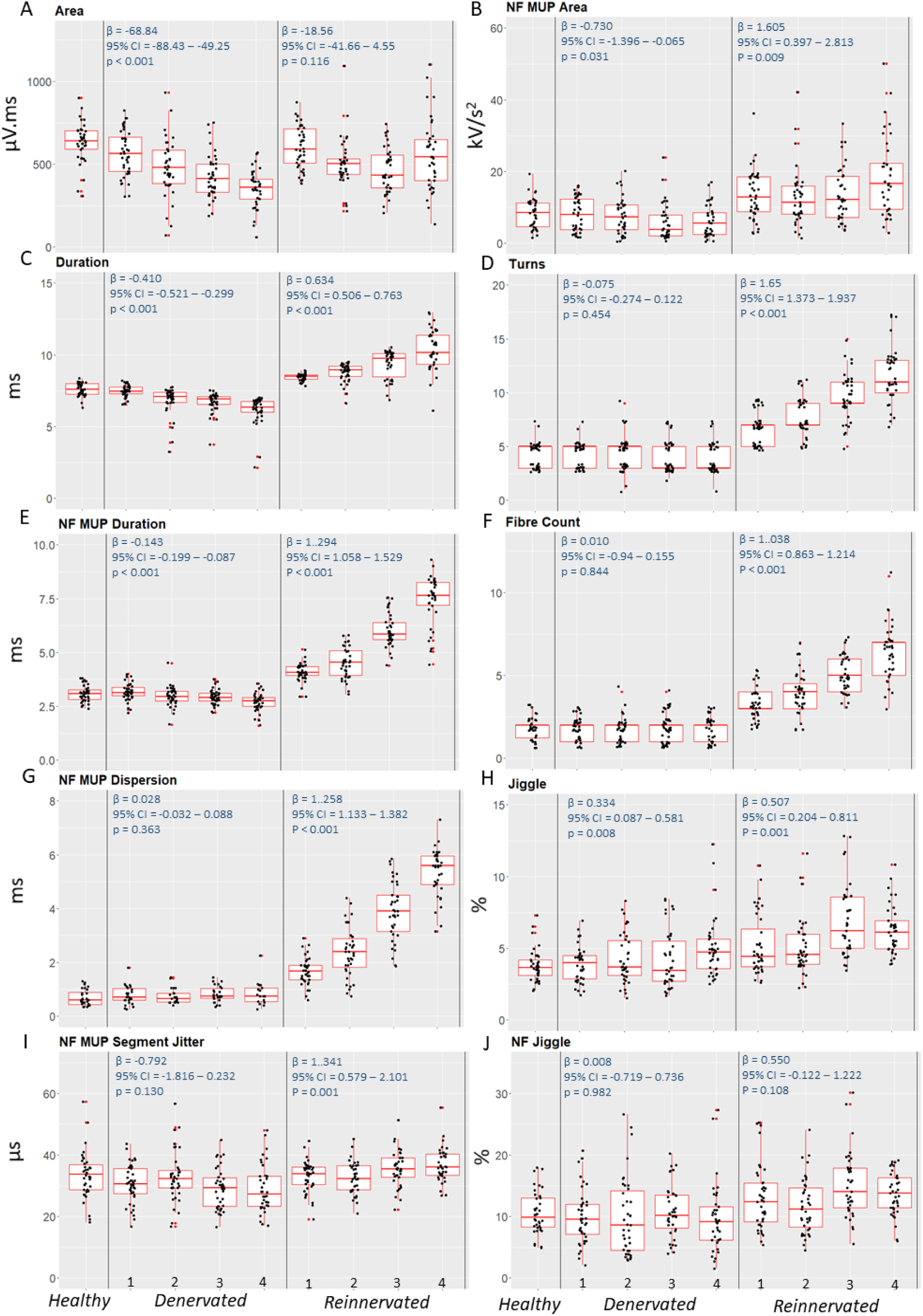
Motor unit potential and near fibre motor unit potential parameters with decreases or increases in the numbers of MU fibres. For increases in the numbers of MU fibres, fibre diameter variability and end plate scatter are also increased (See Table 1 for details). Beta coefficients, 95% confidence intervals and p values from multi level mixed effects linear regression shown inset for each parameter.

Figure 3 panels E and G do not show any trends in NFM duration or NFM dispersion with increased denervation. The constant fibre diameter variability and end plate scatter modelled across the levels of increased denervation modelled resulted in no changes to these NFM measures. These panels do show increasing trends in NFM duration or NF dispersion with increased reinnervation. The increased fibre diameter variability and end plate scatter modelled across the levels of increased reinnervation modelled resulted in clear changes to these NFM measures. Figure 3 panels H, J and I do not show any trends in MUP or NFM jiggle and NFM segment jitter with increased denervation, respectively. These panels do show increasing trends in MUP jiggle and NFM segment jitter, but not NFM jiggle with increased reinnervation. With the constant amount of NMJ jitter modelled across the increasing levels of denervation and reinnervation modelled no trend in instability measures are expected. However, the increased fibre diameter variability and end plate scatter modelled across the levels of increased reinnervation modelled resulted in increased MUP jiggle and NFM segment jitter values.

### 3.4 Increasing levels of jitter (NMJ transmission time variability)

Six levels of jitter (NMJ transmission time variability) were modelled (20, 40, 60, 80, 100, and 120 μs). For each jitter level, ten EMG signals each containing 4 MUPTs were simulated for 10 unique radial needle positions. Forty MUPTs were then extracted from the EMG signals for each level of jitter using DQEMG algorithms. Figure 4 panels A to C show MUP and NFM size parameters across increases in jitter. Increased MFP temporal dispersion associated with increased jitter does not decrease MUP area, except for 120 μs jitter, (Figure 4A), while NFM area (Figure 4B) and MUP duration (Figure 4C) do not change across all jitter values studied. Figure 4 panels D and F show no changes in number of MUP turns or NF count values across all increases in jitter studied. Figure 4 panels E and G show no changes in NFM duration or NF dispersion across all increases in jitter studied. Figure 4 panels H and J show MUP and NFM jiggle values, while panel I shows NFM segment jitter values across increases in jitter. All of these measures clearly reflect the increases in modelled jitter. The temporally-based NFM segment jitter values, with units in μs, had the highest β and relative β values.

**Figure 4.**
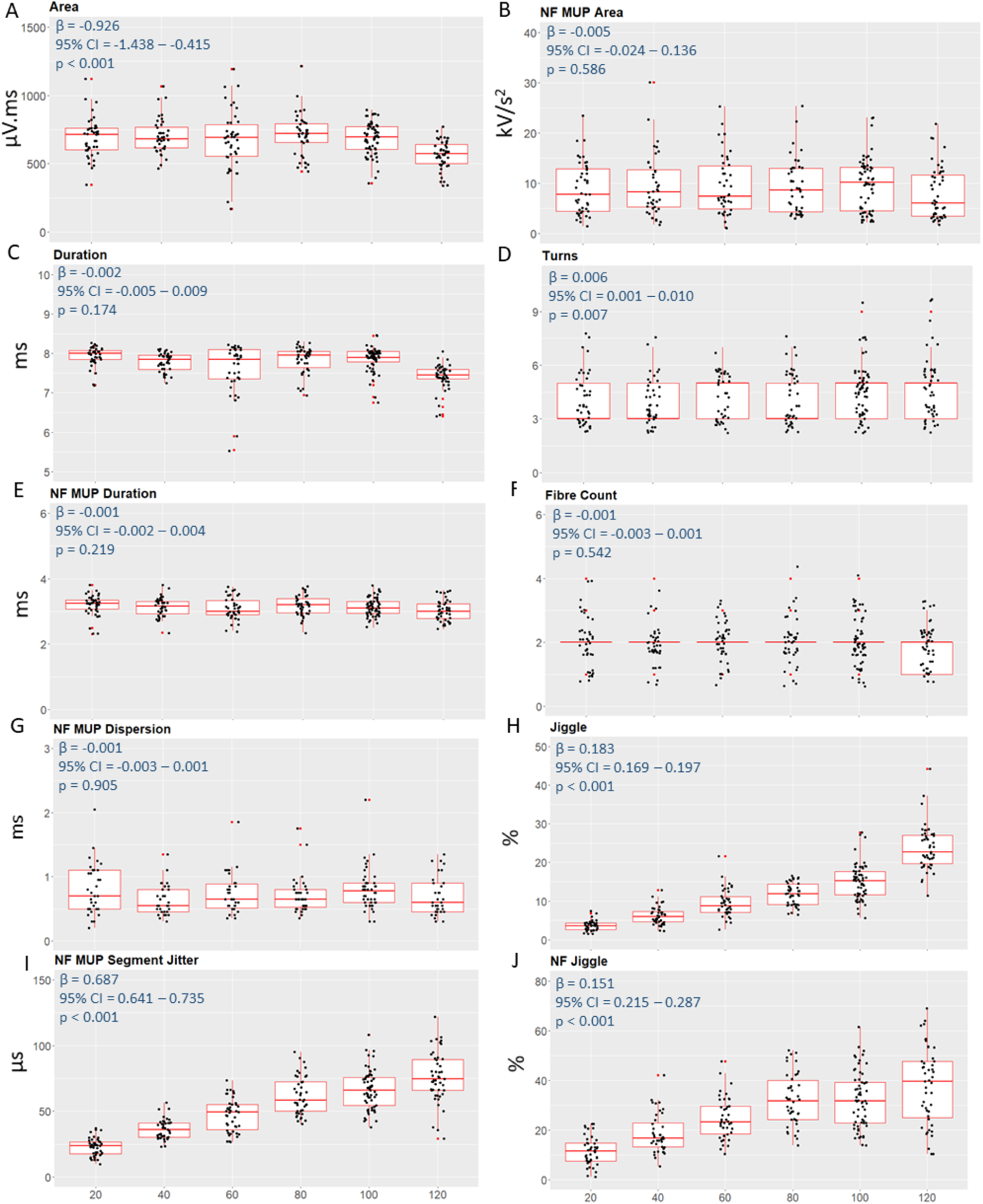
Motor unit potential and near fibre motor unit potential parameters with increased jitter. Beta coefficients, 95% confidence intervals and p values from multi level mixed effects linear regression shown inset for each parameter.

**Figure 5.**
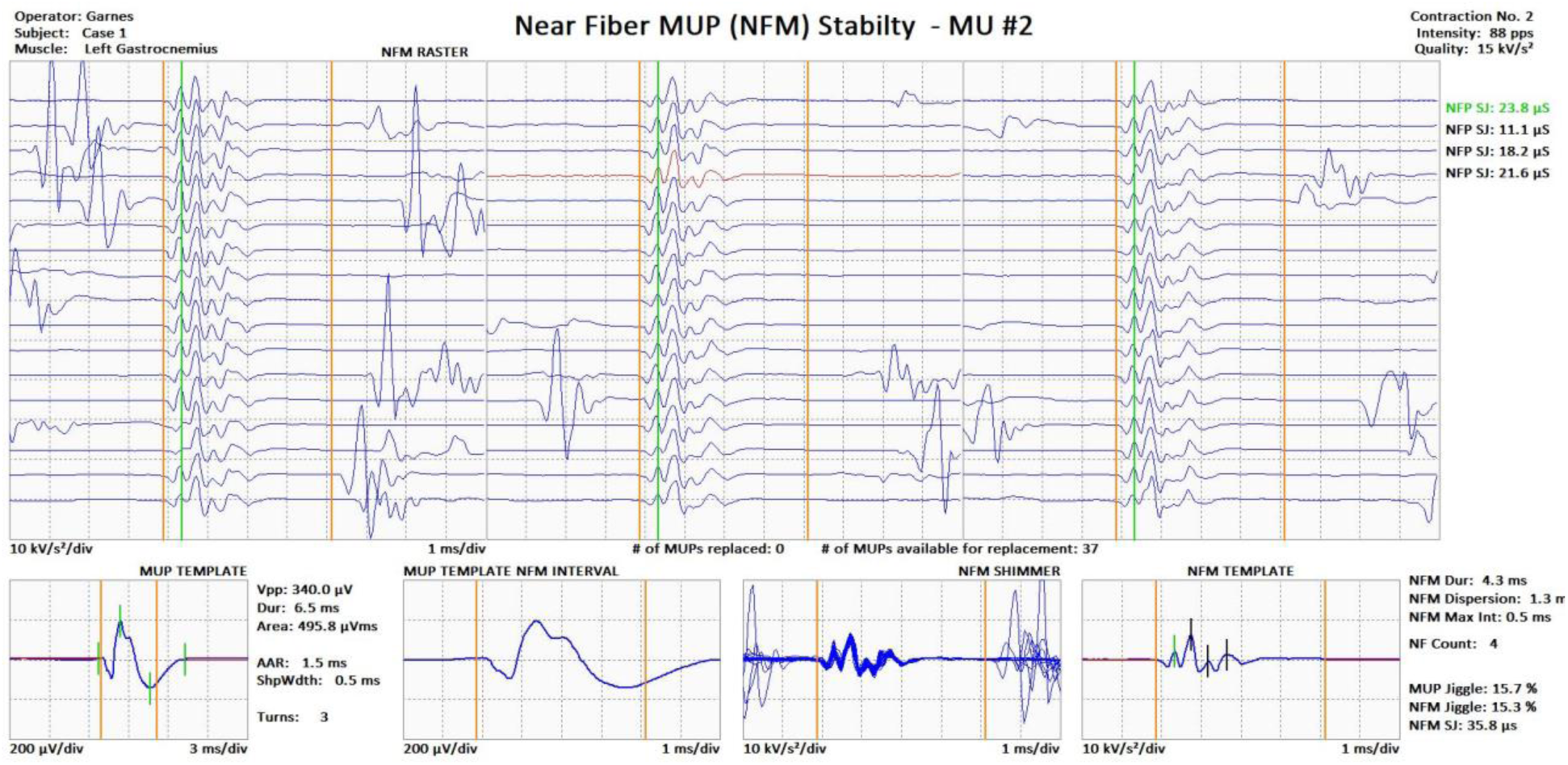
Data from a healty MU in a healty muscle (gastrocnemius). A raster of 3, 12 ms sweep, columns of isolated NFMs, an 18 and 8 ms sweep MUP template, and an 8 ms sweep NFM shimmer and template are shown. The top to bottom vertical lines demarcate the NFM duration, in the NFM template the elements of the NFM count are demarcated by short vertical lines. The interval between the first and last short vertical line is the NFM dispersion. Associated MUP and NFM template as well as MUPT parameter values are shown. The MUP template is of somewhat below average size, the NFM template is of average duration and dispersion and the NFM raster/shimmer is stable. Note that the rasters of isolated NFMs as well as the calculated NFP SJ value indicate healthy NMJ transmission temporal stability in spite of slight needle movement during signal acquisition.

### 3.5 Exemplary Human Patient Data

Figures 5 to 7 show exemplary real EMG data from healthy, enlarging/reinnervating and depleted MUs of a control, neurogenic and myopathic subject, respectively.

Figure 5 shows a normal MUP template, an NFM template with moderate duration and dispersion and a stable NFM raster/shimmer. It corresponds to a 65 y.o. woman presenting with right lumbar pain and occasional paresthesia in the right L5 dermatome. EMG and MRI findings were normal and symmetric compared with the asymptomatic contralateral side. The MUP data displayed was taken from the contralateral asymptomatic gastrocnemius considered as a reference.

Figure 6 corresponds to a 70 y.o. male presenting with progressive asymmetric weakness, muscle mass loss and spasticity over the last year. Needle EMG revealed extended ongoing denervation (i.e. fibrillation potentials and positive sharp waves), fasciculations and reduced recruitment together with polyphasic unstable and enlarged MUPs in three body segments, meeting the revised El Escorial criteria for definite ALS. Panels A to C show different stages of MUP denervation-reinnervation. In panel A, two normal shaped and sized MUP templates along with NFM templates with increased dispersion and moderately unstable NFM shimmers, suggest an early stage of reinnervation. In panel B, two normal sized MUP templates with increased shape complexity as well as NFM templates with increased duration, dispersion and increasingly unstable NFM shimmers suggest an advanced stage of ongoing reinnervation. Panel C displays two enlarged MUP templates associated with NFM templates with increased duration and dispersion, and only moderately unstable NFM shimmers suggesting mature reinnervation, as well as a small MUP template associated with a NFM template with increased duration and dispersion and a stable NFM shimmer, possibly suggesting depletion of an enlarged MUP through degeneration of distal nerve terminals (de Carvalho & Swash, 2016).

**Figure 6.**
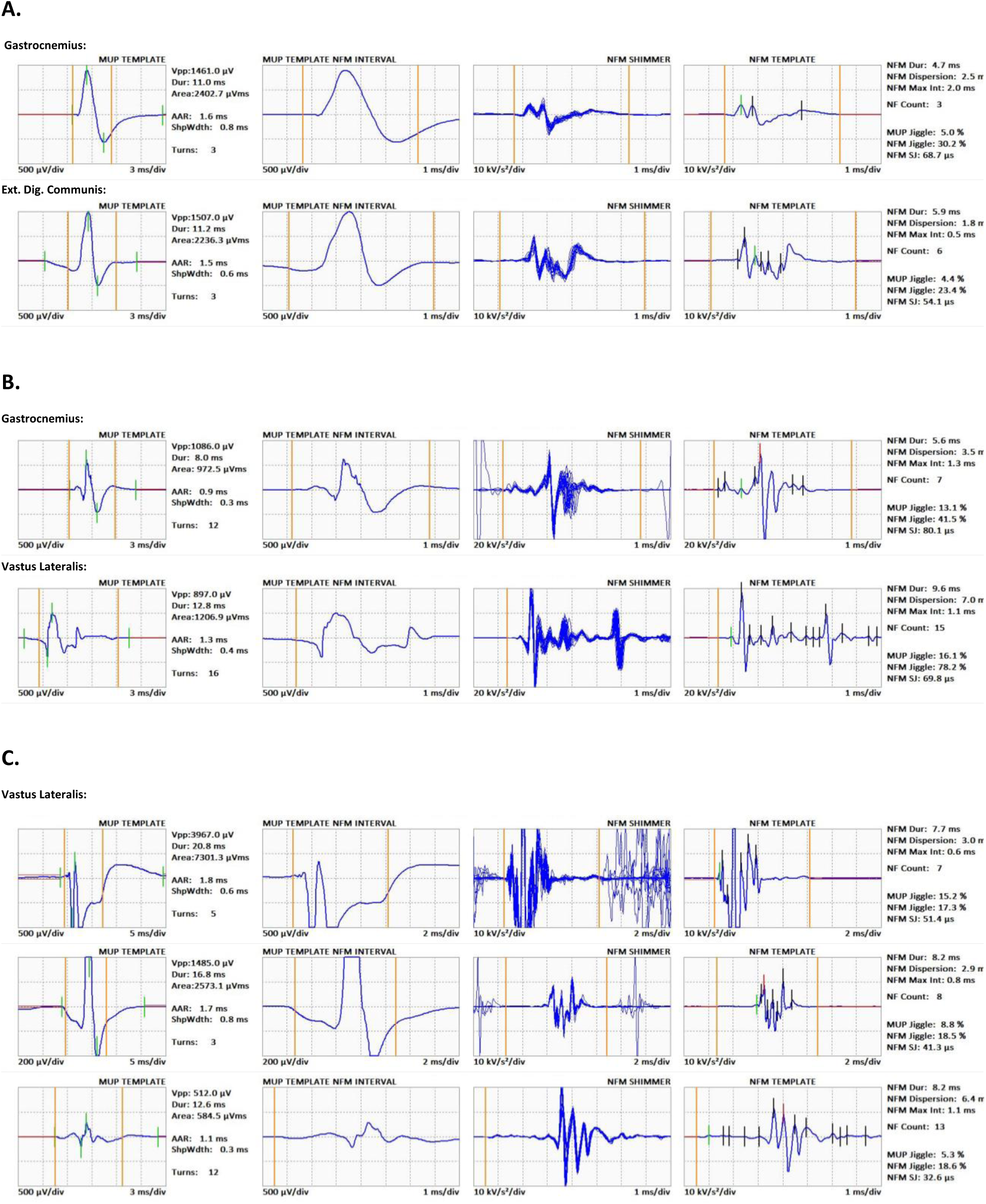
Different stages of reinnervation are depicted across panels A to C. In panel A emerging reinnervation is manifested as increased NFM instability and a mild rise in NFM duration and dispersion. In panel B reinnervation is prominent, showing a complex shaped MUP with increased NFM parameters. In panel C three stable MUPs are depicted. From top to bottom MUP size wanes yet with increased temporal dispersion (high NFM duration and dispersion). It is hypothesised that progression of certain neurogenic disorders can cause partial denervation (see text). From left to right: MUP template, MUP template NFM interval, NFM shimmer and NFM template are displayed. The top to bottom vertical lines demarcate the NFM duration.

Figure 7 corresponds to a middle-aged woman diagnosed with dermatopolymyositis in childhood currently on immunosuppressant at low doses for intercurrent arthritis (Azathioprine), who presented with symmetric residual weakness in proximal muscles during both moderate and sustained efforts. Needle EMG did not show evidence of active myositis and routine nerve conduction studies were anodyne. The figure shows a small size MUP template, complex in shape (increased number of turns) and a NFM template with increased NF counts and dispersion, but duration in the upper limit of normality and a stable NFM shimmer.

**Figure 7.**
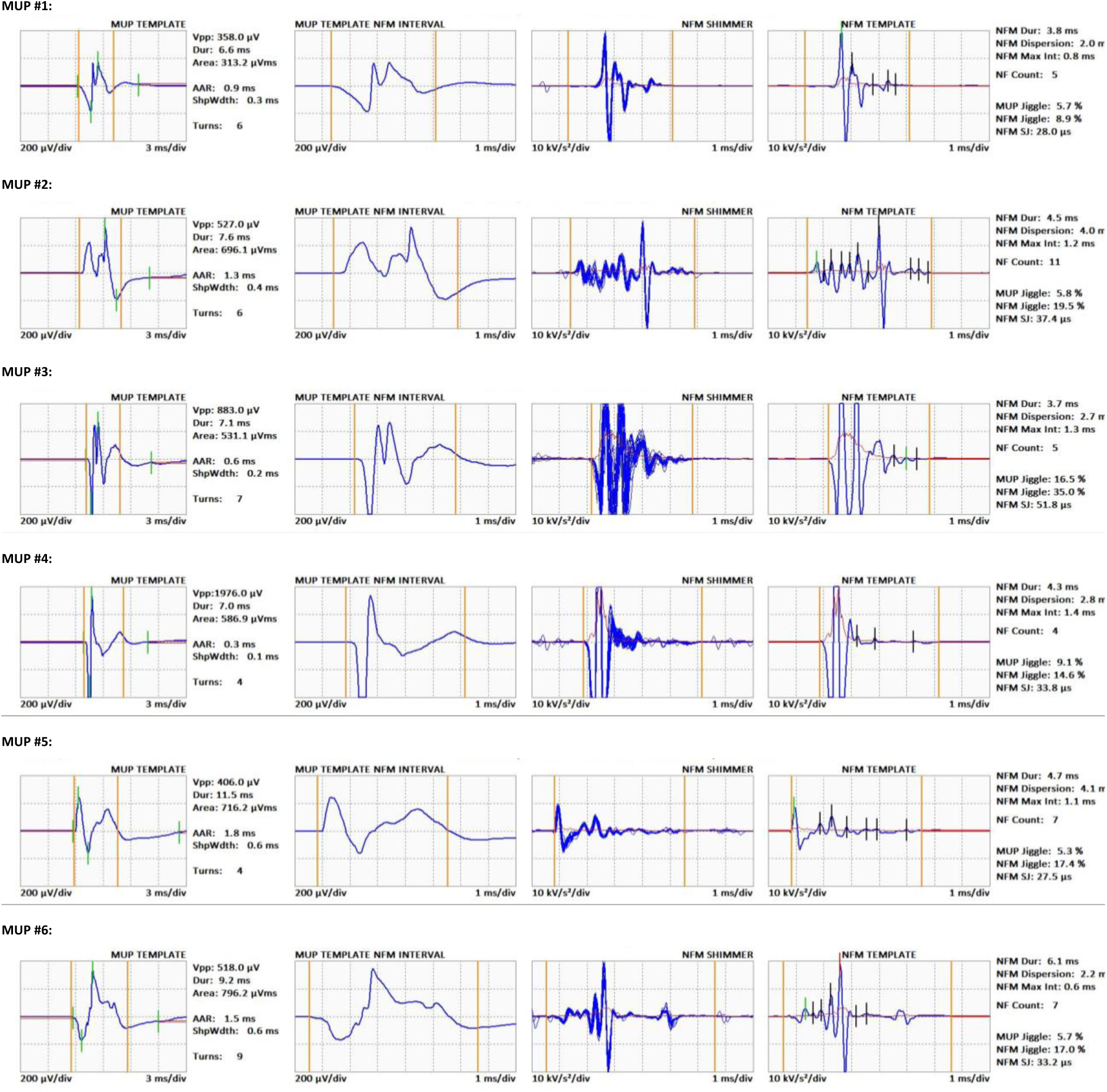
Example of a mild myopathic deltoid showing MUPs of partially depleted MUs recorded during an isometric contraction during low level of activation. Each row corresponds to individual MUs from the same contraction. From left to right: MUP template, MUP template NFM interval, NFM shimmer and NFM template are displayed. The top to bottom vertical lines demarcate the NFM duration.

## 4.0 Discussion

NFEMG focuses on contributions/MFPs of MU muscle fibres lying close to the detection surface of a needle electrode. NFM measurements of the number, temporal dispersion and temporal consistency of generated MFPs are dependent on local MU morphology and electrophysiology and as such can infer subtle MU changes as occurring with ageing and/or disease. As demonstrated with simulations of increased fibre diameter variability or endplate scatter, as the spread in the expected MFAP arrival times (i.e. the times at which the MFAPs are expected to pass closest to the center of the electrode detection surface) increases so do the number and temporal dispersion of detected MFPs. Increasing fibre diameter variability and end plate scatter are early effects of neuromuscular disorders and ageing and precursors to changes in MU size and fibre grouping. Being able to detect these initial MU changes potentially allows early more sensitive detection of MU abnormalities.

The effects of increasing amounts of simulated fibre diameter variability on MUP and NFM template and instability parameters are presented in Figure 1. MUP area decreases with increasing fibre diameter variability (MUP amplitude (V_pp_) follows the same pattern, values not shown) which can be a consequence of increased MFP dispersion associated with increasing fibre diameter variability, which can result in more destructive MFP superpositions causing phase cancelation, even though the number of contributing MFP remains constant. Notably, these effects would likely be reduced with a smaller axial distance separating the electrode and endplate region. MUP duration, in contrast, increased with increasing fibre diameter variability (Figure 1C) which can also be a consequence of increased MFP dispersion associated with increasing fibre diameter variability, ultimately increasing the time interval between MUP onset and end positions. These results are consistent with those of Stålberg & Karlsson (2001) who reported both MUP V_pp_ and area decreases for simulated data with increases in MFP dispersion. NFM area was not affected by increased fibre diameter variability (Fig 1B) which can be related to the relatively smaller number of MFPs that make significant contributions to NFMs. As such, NFM area measures were in general more variable and did not show a clear relationship with fibre diameter variability. Turns and NF count both increased with fibre diameter variability (Fig 1 D, F) without increases in numbers of MU fibres or MU fibre density and specifically due to increases in MFP dispersion allowing increased numbers of MFP contributions to be detected. NFM duration and NF dispersion both increased with increases in fibre diameter variability (Fig 1 E,G). Each of these temporal measures are directly related to MFP dispersion and clearly reflect increases in MFP dispersion caused by increases in the range of MFAP conduction velocities associated with increases in fibre diameter variability. MUP jiggle, NFM jiggle and NFM SJ across MUPTs all had similar modest increases with increased MFP dispersion associated with increased fibre diameter variability, explained by MFP contributions and their associated temporal variability becoming easier to observe.

The results of simulating increasing amounts of end plate scatter on MUP and NFM template and instability parameters are presented in Figure 2. Overall, the respective template parameters were similarly affected by increases in end plate scatter as with increases in fibre diameter variability. However, the effects of the amounts of endplate scatter were greater than those of the amounts of fibre diameter variability. The levels of endplate scatter simulated generated greater amounts of MFP dispersion which is reflected in the increased trends of the template parameter values. The MUP and NFM instability measures all had similar modest increases, which were, despite the increased amounts of MFP dispersion, similar in extent as with the fibre diameter variability simulations (Fig 2 H,J,I).

The effects of simulating MU remodelling, the denervation and reinnervation of fibres, on MUP and NFM template and instability parameters are presented in Figure 3. These data demonstrate effects of decreasing numbers of MU fibres (denervation), and increasing numbers of MU fibres along with increasing fibre diameter variability and endplate scatter (reinnervation). For MUs with fewer numbers of fibres, MUP area and duration decreased with decreasing numbers of MU fibres. The reversal of this pattern could be assumed of all other modelled parameters remained constant. For MUs with greater fibre numbers, diameter and endplate variability, MUP area clearly increased only after 40 fibres were added, while duration consistently increased with increasing numbers of MU fibres (Fig 3 A, C). For both scenarios NFM area remained constant but became more variable with increasing numbers of MU fibres and the increasing MFP dispersion modelled (Fig 3 B). Turns and NF count remained constant across decreasing numbers of MU fibres and both increased with increasing numbers of MU fibres and the increasing MFP dispersion modelled (Fig 3 D, F). NFM duration and dispersion remained constant across decreasing numbers of MU fibres and both increased with increasing numbers of MU fibres and the increasing MFP dispersion modelled (Fig 3 E, G). Across the increasing numbers of MU fibres simulated, NFM duration and dispersion clearly reflect the increasing level of MFP dispersion modelled. Despite the increased amounts of MFP dispersion modelled, the MUP and NFM instability measures all had similar modest increases, which were similar in extent as with the fibre diameter variability simulations (Fig 3 H,J,I).

With or without MFP dispersion, MUP V_pp_ and area values were strongly correlated (r=0.82, data not shown). Without MFP dispersion (denervation), MUP area and MUP duration decreased and reflects the number of MU fibres. However with MFP dispersion (reinnervation), MUP area showed little change and MUP duration increased, therefore either may not reflect the number of MU fibres. NFM area is less affected by MFP dispersion than MUP area and duration. Therefore, if MFP dispersion is increased, NFM area may not be strongly correlated with MUP area or duration but nonetheless can be reflective of the number of MU fibres. NF count is well correlated with number of turns and can equally reflect the MFP dispersion underlying MUP/NFM complexity. NF count is not directly related to fibre density measurements obtained using a SFEMG electrode (Stalberg and Thiele 1975) because to obtain the latter measurements, the needle is positioned to obtain minimal MUP rise times from a single MU, whereas for the former, the needle is positioned to obtain suitably sharp MUPs across the interference pattern. NFM duration is strongly correlated with NFM dispersion and both most clearly and directly reflect MFP dispersion.

The results of simulating increasing amounts of jitter on MUP and NFM template and instability parameters are presented in Figure 4. MUP area and duration drop for high jitter values (Fig 4 A, C). All other MUP and NFM template parameters are unaffected by increasing jitter. However, MUP and NFM jiggle and NFM SJ values increased with increasing jitter (Fig 4 H, J, I) and are therefore all strongly correlated with and clearly reflect NMJ transmission time variability.

Unlike fibre pair jitter, measured using a single-fibre or concentric needle electrode to detect sets of MFP contributions from isolated pairs of muscle fibres (Sanders & Stålberg, 1996; Stålberg & Sanders, 2009), MUP and NFM jiggle and NFM SJ represent the amount of MFP temporal variability across all of the MFPs significantly contributing to the MUPs/NFMs of a MUPT. Jiggle expresses MFP temporal variability (primarily NMJ transmission time variability) by measuring MUP instability along the amplitude axis. Jiggle, measured using sums of median consecutive absolute amplitude differences, which reduce the effects of small needle movements and contamination from other MUs, and the ratio to template area, which reduces the effects of needle focusing/positioning, was determined the best of several instability measures considered by Stålberg and Sonoo, 1994. Nonetheless, in real signals, MUP jiggle values are more contaminated by contributions from other MUs (Campos et al. 2000) than are NFM jiggle or NFM SJ values. Algorithms used in this work to extract MUPTs and to select isolated MUPs/NFMs are specifically designed to account for MUP/NFM instability, to minimize the effects of needle positioning/focusing and to effectively align MUPs/NFMs for instability assessment to reduce contamination and other sources of instability measurement error described in earlier works (Campos et al., 2000; Stålberg & Sonoo, 1994).

With jitter held at a constant “reference” value (25 μs), MUP and NFM jiggle and NFM SJ increased with MFP temporal dispersion as the effects of more individual MFP components could be determined (Fig. 1 H,I,J and Fig. 2 H,I,J). However, the increased values do not exceed “reference” values (<50%, < 50 μs). Stålberg & Sonoo (1994) discussed how MUP polyphasia, caused by increased MFP temporal dispersion exposes inherent/true instability which can be masked with decreased MFP temporal dispersion where jittering MFPs are simply consistently superimposed such that the apparent/measurable jitter/temporal instability is reduced. In other words, instability measures can increase due solely to MFP temporal dispersion as the inherent/true temporal variability can be more accurately measured. This in turn suggests that increased polyphasia without increased jiggle can indicate chronic myopathic fibre diameter variability increases (Stålberg & Sonoo, 1994). MUP jiggle was affected more by mean MFP temporal dispersion than were NFM jiggle and NFM SJ and NFM SJ was affected more by mean MFP temporal dispersion than was NFM jiggle. Due to the larger MUs simulated, 100^+^ here versus their 10 fibres, the drop in MUP jiggle versus MU size reported by Stålberg & Sonoo was not apparent in this data for either MUP and NFM jiggle of NFM SJ (Fig 3 H,J,I). Even though for small amounts of jitter (20 μs), its mean values were relatively high and for high amounts of jitter (100 and 120 μs) its mean values were relatively low, the temporally based NFM SJ measure had the strongest relationship with increasing jitter. Because increases in NFM SJ more closely track those of simulated jitter it can be considered more directly related to the inherent temporal instability of MFP contributions to MUPs than are MUP and NFM jiggle, and is therefore more sensitive for detecting temporal instability in MUPTs.

Figures 5 to 7 show exemplary real data recorded from healthy, expanding and depleted MUs, respectively. Results from a healthy MU (Fig 5) show a moderate MUP area (size), with average NFM duration and dispersion and stable NFM raster and shimmer plots (healthy NMJ transmission). Results from the expanding MU (Fig 6) show two average size MUPs (panel A), with incipient increases in NFM duration and dispersion and increased instability in the NFM shimmer plot (increased NMJ transmission time variability reflecting emerging reinnervation). In panel 6B results from two average size MUPs with increased shape complexity show prominent increases in NFM duration, dispersion and NFM shimmer instability, reflecting prominent reinnervation. Whereas panel 6C shows an enlarged, a moderately enlarged and a reduced sized MUP, all with increased and similar NFM duration and dispersion values and rather stable NFM shimmer plots, possibly resembling distal axonal damage affecting longstanding enlarged and stable MUs as part of the neurogenic process. Results from the depleted MU (Fig 7) show slightly reduced MUP sizes, with increased NFM duration and dispersion and stable NFM shimmer plots.

The NFM duration and dispersion (7.7, 8.2 and 3.0, 2.9 ms, respectively) measured for the two expanding MUs in Figure 6C are clearly longer than for the healthy MU (4.3 and 1.3 ms, respectively) and greater than for any values obtained from simulations of only increased fibre diameter variability. This suggests, an MFP dispersion associated with more than just increased fibre diameter variability. It suggests some degree of increased end plate scatter and possibly some increased fibre diameter variability consistent with axonal branch sprouting and possibly some fibre atrophy and/or hypertrophy. The increased NFM SJ value in both MUPs in panel 6A (68.7 and 54.1 μs) clearly suggests increased NMJ transmission time variability consistent with immature/nascent NMJ formations. All of these NFM results are consistent with ongoing reinnervation and given that their respective MUP areas (2402 and 2236 μVms) are only slightly increased and duration is within the normal range (11.0 and 11.2 ms, respectively) suggests an initial/early stage of reinnervation.

When considering myopathic depleted MUs (Fig 7), NF count, NFM duration and dispersion values, for all the MUs sampled from a single contraction, are longer than for the healthy MU (4, 4.3 ms and 1.3 ms, respectively) and greater than for any values obtained from simulations of only increased fibre diameter variability. This suggests an MFP dispersion associated with more than just increased fibre diameter variability. It suggests some degree of increased end plate scatter and possibly some increased fibre diameter variability consistent with axonal branch sprouting and possibly some fibre atrophy and/or hypertrophy. The reduced MUP areas and normal durations suggest MUs may have reduced numbers of fibres of variable diameter and possibly some increased end plate scatter.

Sonoo, 2002 suggests the most important aspects of MU changes caused by neuromuscular disorders manifest as recruitment abnormalities, rather than as changes in MUP morphology. However, NFM measurements have the ability to reflect MU changes before the MUs and/or the numbers of MUs in a muscle significantly change and therefore before significant changes in MUP size (See Fig. 6 B). Sonoo, 2002 also suggests that needle position (focusing) and the level of muscle activation during MU sampling, greatly influence the parameters of sampled MUPs. NFM measures of dispersion and instability are not directly dependent of MUP size and shape and therefore are less affected by focusing and level of activation. And finally, Sonoo, 2002 suggests that MUP duration, considered to be the cardinal parameter in MUP analysis, has several drawbacks, including high measurement variability and low discriminant sensitivity. This is no doubt true when baseline noise affects onset and end marker positions. However, for low noise waveforms, as with the noise-free simulated data, MUP duration, as suggested by (Buchthal and Rosenfalck 1955), is most sensitive to MU size. MUP duration increases with MU size and MFP dispersion, while MUP area increases with MU size but decreases with MFP dispersion, which often accompanies MU enlargement via reinnervation, and therefore confounds the relationship between MU size and MUP area (See Figs. 1C, 2C, 3C and 4C). Nonetheless in practice, low-noise waveforms may not be available and therefore area, which is highly repeatable in young and old muscles (Piasecki et al. 2018), and less affected by specific onset and end marker positions may better reflect MU size, despite its decreases with MFP dispersion. Actually, increased MUP duration results from several factors in addition to the number of innervated muscle fibres, including end-plate scatter, NMJ transmission delay, and slow conduction through terminal branching axonal sprouts. This results in decreased synchronicity of muscle fibre activation (i.e. MFP generation) across a motor unit, and increased MUP duration (de Carvalho & Swash, 2016). As shown in the simulated data results, NFM duration and NF dispersion are both very sensitive to MFP temporal dispersion caused by fibre diameter variability and/or end plate scatter (which as simulated can also account for NMJ transmission delay and slow axonal branch conduction).

With active denervation and reinnervation MUPs and NFMs are unstable. Newly-formed, immature end-plates have immature acetylcholine receptor subunits, with a lower safety factor for neuromuscular transmission (Stålberg, 1982; Stålberg & Antoni, 1980). In addition, slowed distal motor nerve conduction in partially myelinated regenerating nerve fibres and atrophied, newly reinnervated muscle fibres leads to increased fibre diameter variability and end plate scatter which results in sufficient MFP dispersion that the inherent NMJ transmission time variability can be accurately measured. For these measurements, Stålberg and Sonoo (1994) suggest MUP instability, the ‘jiggle’, is best evaluated using a 500 Hz or1 kHz high-pass filter, giving important qualitative information regarding MUP morphology and MUP instability. NFM jiggle and NFM SJ are both based on high-pass filtered signals and both can provide useful measures strongly related to NMJ transmission time variability. MUP instability anticipates the development of neurogenic MUPs as shown by concentric needle EMG (de Carvalho & Swash, 2013) and can be useful in differential diagnoses. (See Fig 6A and B). Furthermore, de Carvalho et al., 2014 report that mean MUP duration and jitter increases are the most sensitive parameters for tracking ALS and that at the end stages of ALS, small MUPs may be observed, possibly resulting from degeneration of distal nerve terminals through axonal dying-back pathology (de Carvalho & Swash, 2016) (possibly the lower MU shown in Fig 6C). As such, MUP area, NFM duration, NFM dispersion, NFM jitter and NFM SJ, assessed together are well suited for detecting and tracking MU morphology and electrophysiological changes.

## Conclusion

Standard needle EMG examination can detect fibre denervation, reinnervation, loss, atrophy and hypertrophy, but detecting initial changes and quantifying their extent and progress requires more sophisticated techniques which can also provide a temporal profile of a disease process across longitudinal examinations. The data presented here demonstrate NFEMG methods can augment a standard needle EMG examination with quantitative MU morphological and electrophysiological information and can be applied to study the neural effects of specific disease processes and ageing.

## Data Availability

The datasets generated and analysed during the current study are available from the corresponding author upon reasonable request.

## Acknowledgements

Mathew Piasecki is supported by the Medical Research Council [grant number MR/P021220/1] as part of the MRC-Versus Arthritis Centre for Musculoskeletal Ageing Research awarded to the Universities of Nottingham and Birmingham, and by the NIHR Nottingham Biomedical Research Centre.

